# Autonomous generation of decision-grade clinical evidence

**DOI:** 10.64898/2026.06.26.26356653

**Authors:** Shaopeng Yang, Jiayu Wu, Hao Xie, Zhuoyao Xin, Wei Wang

**Author notes:** These authors contributed equally (Shaopeng Yang, Jiayu Wu, Hao Xie). **Corresponding authors** Wei Wang, MD, PhD, State Key Laboratory of Ophthalmology, Zhongshan, Ophthalmic Center, Sun Yat-sen University, Guangzhou, Guangdong, China.

## Abstract

Medical practice is bottlenecked by the slow production of high-quality clinical evidence. Despite progress in automating selected stages, autonomous conduct of the entire research life cycle remains beyond reach. Here we present OpenEBM, the first autonomous system to generate decision-grade clinical evidence by conducting evidence-synthesis research end to end. To enable and evaluate this, we develop OpenEBM-Corpus, a foundation resource of expert-annotated research trajectories that enables training of a specialist model, and OpenEBM-Bench, a multidisciplinary benchmark that evaluates the entire research life cycle. Our compact specialist model generates valid clinical evidence in 90.7% of end-to-end evaluations and matches expert performance across the research trajectory, whereas GPT-5 falls to 3.8% as failures propagate through dependent stages. In blinded evaluations across clinical domains, independent evaluators prefer OpenEBM at multiple stages and cannot distinguish its reasoning traces from expert-conducted work above chance. Applied to a question left unresolved by current guidelines, OpenEBM produces *de novo* evidence addressing the efficacy and safety of neoadjuvant chemotherapy for locally advanced rectal cancer. OpenEBM brings within reach the founding aspiration of evidence-based medicine and establishes a paradigm for scalable evidence generation.

## Main

For most of medical history, clinical decisions were governed by experience and local judgement. Evidence-based medicine (EBM) broke with this epistemology by requiring clinical claims to rest on empirical evidence.^1, 2^ At the center of this transformation is evidence synthesis, a distinctive form of research that operates above primary trials to generate decision-grade clinical evidence from dispersed, fallible observations.^3, 4^ Yet the research process that made medicine evidence-based has itself become a bottleneck.^5, 6^ Each body of evidence it produces demands years of expert labor and direct costs exceeding US$140,000, while novel therapies, emerging public-health threats, and expanding literature continually raise urgent questions that outpace human research capacity.^6–8^ Existing evidence quickly becomes outdated, and major gaps persist for rare diseases, low-resource settings, and under-represented populations.

Large language models (LLMs) accelerate access to biomedical knowledge and achieve strong performance in question answering and narrative literature synthesis.^9–11^ However, clinical evidence generation requires execution of the complete EBM research trajectory, in which protocol-bound decisions across dependent stages determine the evidentiary data and the conclusions they justify. In our experiments, state-of-the-art LLMs missed 66.1–75.5% of trial data, misjudged risk of bias in 68.6–83.0% of cases, and generated valid evidence in only 3.8% of end-to-end attempts. These failures reveal a fundamental mismatch between models trained to reproduce existing knowledge and the capabilities required to execute EBM research trajectories. Moreover, evaluations are equally limited, confined to narrow clinical domains or isolated subtasks rather than the full research life cycle.

We introduce OpenEBM (**Figs. 1a–b**), to our knowledge the first autonomous system to produce decision-grade clinical evidence by executing the full life cycle of evidence-synthesis research. OpenEBM unifies three advances: (1) a domain-specialized corpus (OpenEBM-Corpus) with expert-annotated EBM research trajectories; (2) a compact specialist model (OpenEBM-8B) trained to conduct EBM research; and (3) an inference pipeline whose protocol-grounded generation constrains autonomous research within explicit methodological rules. OpenEBM-Corpus provides an up-to-date resource of 5.7 million EBM research trajectories curated from real-world expert practice, enabling a compact model to learn EBM research conduct without reliance on proprietary LLMs. These advances equip an openly trained model to produce end-to-end clinical evidence that remains beyond reach even for frontier systems.

**Fig. 1.**
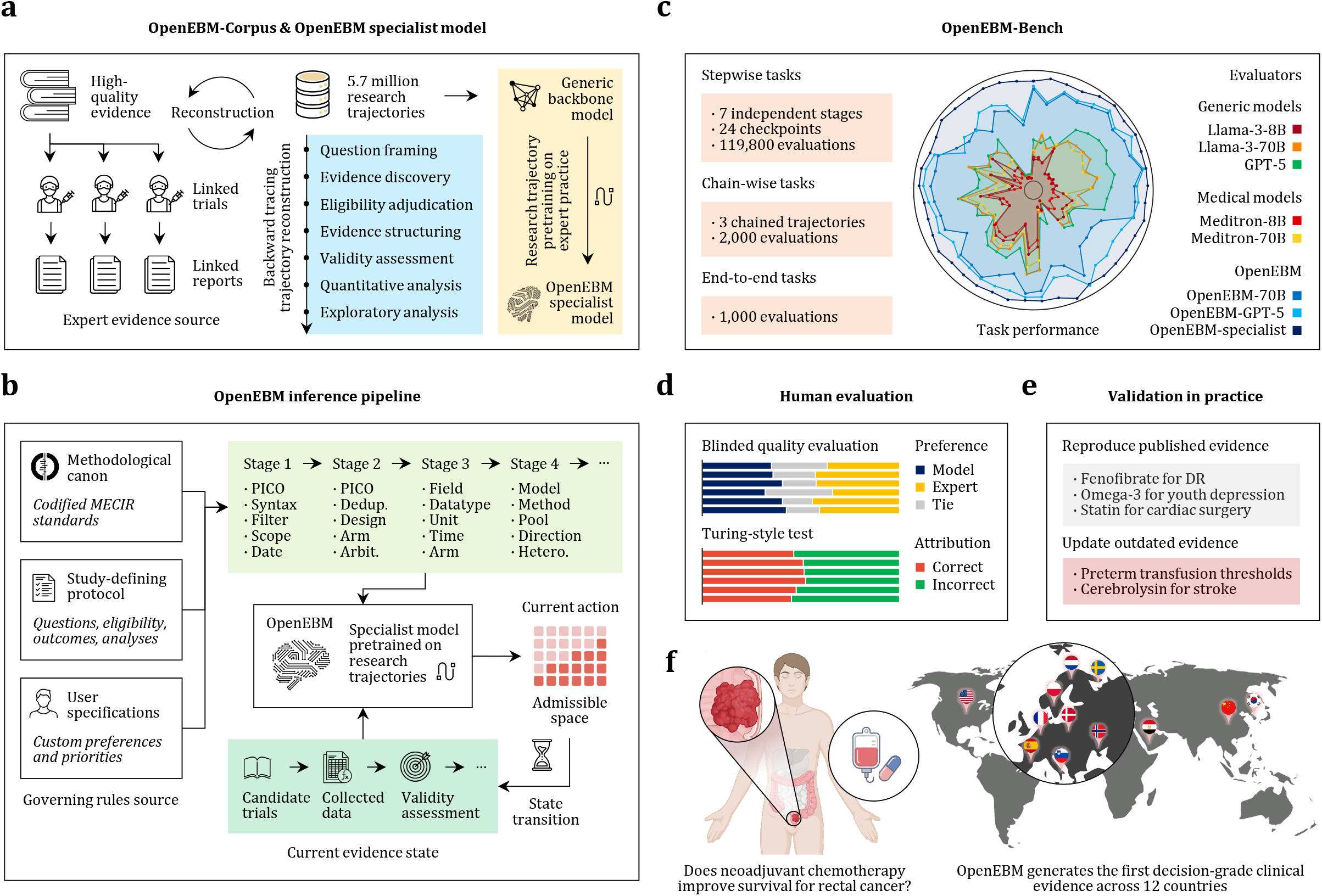
Overview of OpenEBM for autonomous evidence-synthesis research. (**a**), Construction of OpenEBM-Corpus and training of the OpenEBM specialist model. Published expert evidence syntheses are linked to their underlying trials and reports, then reverse-reconstructed into EBM research trajectories that supervise a compact specialist model. (**b**), OpenEBM inference pipeline. Methodological canon, study-defining protocols, and user specifications are resolved into governing rules that constrain each stage of the research trajectory. The specialist model selects admissible research actions, updates the evolving evidence state, and advances the study through protocol-grounded generation. (**c**), OpenEBM-Bench. The benchmark evaluates the entire life cycle of the research process across stepwise, chain-wise, and end-to-end tasks. Performance is compared across generic models, medical models, and OpenEBM systems. (**d**), Human evaluation. Independent experts assess OpenEBM outputs against expert-authored outputs in blinded quality evaluation and Turing-style source-identification tests. (**e**), Validation in practice. OpenEBM is evaluated in real-world evidence-synthesis settings by reproducing published evidence and updating outdated evidence from established studies. (**f**), OpenEBM conducts the first evidence-synthesis study addressing whether neoadjuvant chemotherapy improves survival in locally advanced rectal cancer, integrating randomized trial evidence across 12 countries to generate decision-grade clinical evidence.

To evaluate OpenEBM, we introduce OpenEBM-Bench (**Figs. 1c–d**), a multidisciplinary benchmark for autonomous clinical evidence generation. OpenEBM-Bench captures the full research life cycle of evidence synthesis through 24 major action checkpoints and 15 chained or end-to-end execution trajectories spanning 33 medical and health domains under strict leakage controls. In total, it comprises 122,800 evaluations grounded in real-world research practice. We evaluated state-of-the-art proprietary and open-source LLMs, including GPT-5, Llama 8B and 70B, with and without the OpenEBM inference pipeline. Although GPT-5 outperformed smaller models, any backbone paired with the OpenEBM pipeline consistently surpassed GPT-5 alone. The strongest performance came from our specialist model trained on EBM research trajectories, which sustained end-to-end execution at a fraction of the scale and cost.

Beyond automated benchmarking, we conducted a blinded expert evaluation of 50 held-out studies performed by OpenEBM across five domains (**Fig. 1e**). Independent evaluators assessed OpenEBM against multidisciplinary expert teams whose studies constitute the methodological gold standard for clinical evidence generation, yielding 19,500 fine-grained judgements spanning the full research trajectories. OpenEBM matched the reliability and evidentiary quality of these expert teams. Although the two reasoned differently, neither held a consistent quality advantage. At the decisive stage of issuing a clinically consequential conclusion, evaluators more often preferred OpenEBM for its completeness and reasoning quality. In a Turing-style source-identification test, accuracy stayed at chance across all output categories; even the most confident attributions provided no reliable signal separating OpenEBM from expert work.

Finally, we demonstrate OpenEBM’s capacity to reproduce, update, and—where no prior answer exists—generate clinical evidence *de novo*. OpenEBM investigated the efficacy and safety of neoadjuvant chemotherapy for locally advanced rectal cancer, addressing a major unresolved question in current guidelines that recommend this treatment based on surrogate endpoints without consistent evidence for survival benefit.^12–14^ OpenEBM produced high-certainty evidence under GRADE that neoadjuvant chemotherapy improved overall survival compared with radiotherapy alone and remained superior to adjuvant chemotherapy, with the clinical validity of the findings independently corroborated by a multidisciplinary expert panel. Beyond the scope of current guidelines, OpenEBM further identified a serious-adverse-event signal alongside the benefit. These findings illustrate the potential of autonomous evidence generation to supply missing evidence and refine clinical decisions.

### OpenEBM achieves autonomous evidence generation on OpenEBM-Bench

We evaluated autonomous clinical evidence generation using OpenEBM-Bench, a trajectory-annotated benchmark of 122,800 fine-grained evaluations across 33 disciplinary domains, constructed under strict leakage controls (**Methods**). The benchmark assesses methodological decision-making and protocol-grounded conduct across the full research life cycle using task-specific criteria (**Methods**). To separate the contributions of base-model capability, protocol-grounded inference, and EBM research-trajectory pretraining, we compared three classes of systems: (1) generic and medical LLMs (GPT-5, Llama-3-8B/70B, Meditron-8B/70B); (2) the OpenEBM inference pipeline coupled to off-the-shelf GPT-5 or Llama-3-70B back ends (OpenEBM-GPT-5, OpenEBM-70B); and (3) OpenEBM-8B, which integrates the OpenEBM inference pipeline with a specialist model pretrained on EBM research trajectories.

#### Stepwise evaluation

Generic and medical LLMs showed systematic failures in stages requiring protocol adherence, structured reasoning, and methodologically constrained judgement (**Figs. 2a–d**). In trial ascertainment, strategies generated by GPT-5, Llama-3-70B, and Llama-3-8B failed to retrieve 38.9–89.3% of trials. The same models made incorrect eligibility adjudication for 34.8–45.9% of trials, failed to collect 66.1–75.5% of arm-level datapoints, and produced incorrect risk-of-bias assessments in 68.6–83.0% of cases. The full stepwise performance profile of these models is summarized in **Supplementary Table S1**. Although larger models showed partial improvement over smaller models, scale alone is unlikely to resolve these failures. Continued pretraining on generic medical corpora, as represented by Meditron-8B/70B, yielded no consistent gains over the base models.

**Fig. 2.**
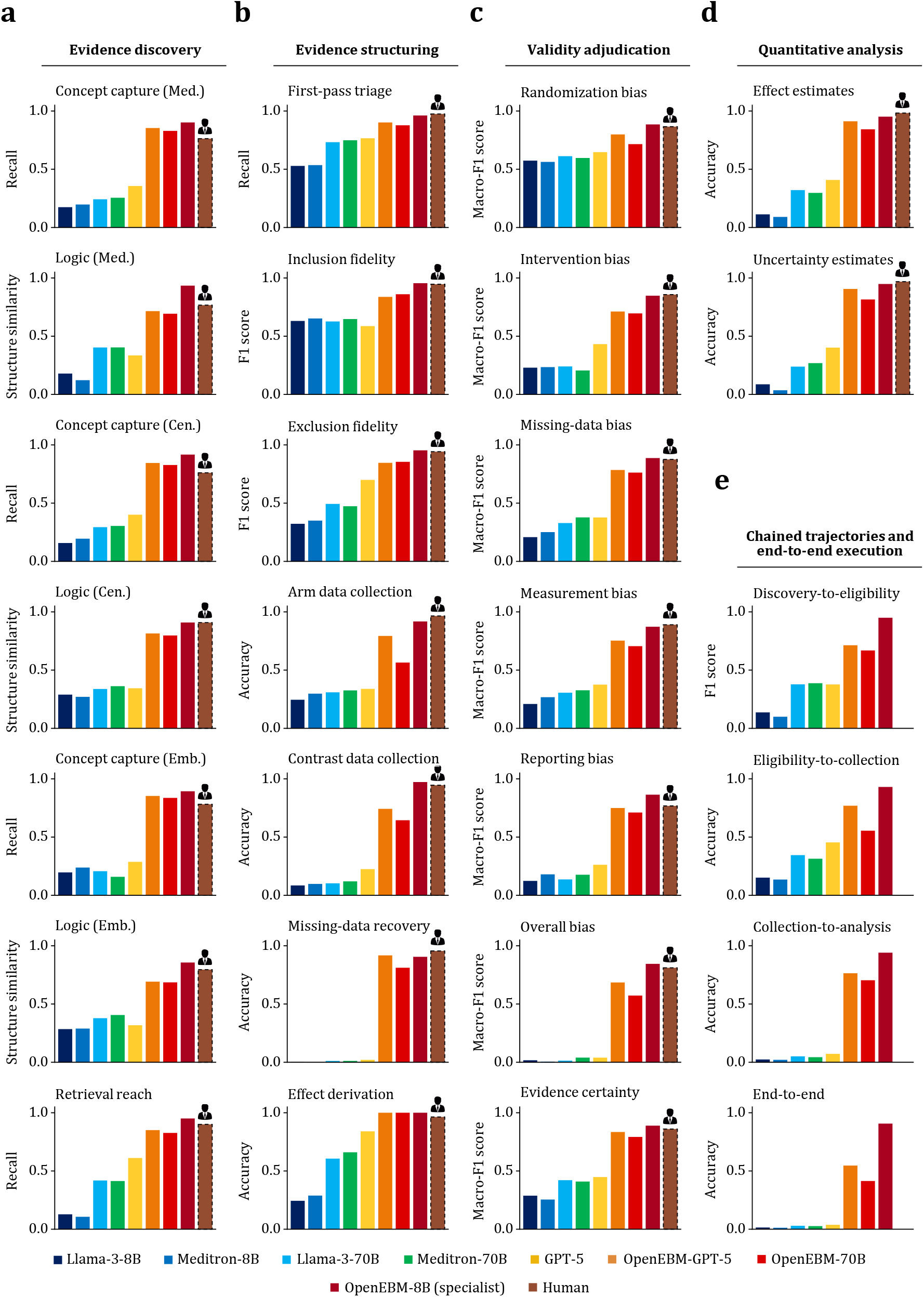
OpenEBM-Bench performance across the research life cycle. (**a**), Trial ascertainment. Performance across concept capture and Boolean logic structure across MEDLINE, CENTRAL and Embase, and overall retrieval reach. (**b**), Eligibility adjudication and data collection. Performance across first-pass triage, inclusion and exclusion fidelity, arm-level and contrast-level data collection, missing-data recovery, and effect derivation. (**c**), Risk-of-bias assessment and certainty-of-evidence adjudication. Macro-F1 scores across bias domains (randomization process, intended interventions, missing outcome data, measurement of the outcome, and selection of the reported result) and evidence certainty. (**d**), Quantitative analysis. Accuracy of effect estimates and uncertainty estimates. (**e**), Chain-wise and end-to-end execution. Performance when outputs are carried forward through dependent stages, including discovery-to-eligibility, eligibility-to-extraction, extraction-to-synthesis and full end-to-end execution. Human experts were evaluated only in stepwise tasks and did not participate in chain-wise or end-to-end execution.

OpenEBM consistently outperformed other systems across stepwise evaluations (**Figs. 2a–d**). Against Llama-3-8B and Llama-3-70B, OpenEBM-8B and OpenEBM-70B improved performance by 7.4- and 2.0-fold in trial ascertainment, 1.8- and 1.5-fold in eligibility adjudication, 3.7- and 1.8-fold in data collection, and 5.0- and 4.1-fold in risk-of-bias assessment. OpenEBM-70B outperformed GPT-5 on most stepwise evaluations, suggesting that protocol-grounded inference improves model outputs by constraining generation within structured EBM research trajectories; applying the same pipeline to GPT-5 improved performance further. OpenEBM-8B achieved the strongest overall performance, surpassing OpenEBM-GPT-5 across stepwise evaluations despite its far smaller backbone, which isolates a gain from EBM-trajectory pretraining beyond model scale or protocol-grounded inference alone.

#### Full research trajectories from protocol to clinical evidence

Because failures propagate across dependent stages, OpenEBM-Bench next evaluated three chained trajectories within the research process (**Methods**). In these chain-wise evaluations, outputs from upstream stages were carried forward as inputs to downstream stages to test whether systems preserve performance across sequential research conduct rather than isolated checkpoints. As expected, all systems performed worse chain-wise than stepwise, indicating error propagation along the trajectory (**Fig. 2e**). This degradation was most pronounced for generic and medical LLMs, whose performance approached zero in chains involving data collection and quantitative analysis. By contrast, OpenEBM-70B and OpenEBM-GPT-5 retained substantially stronger performance across chained trajectories, and OpenEBM-8B achieved the best performance.

OpenEBM-Bench next evaluated whether systems could complete the full research life cycle without access to any gold intermediate outputs (**Methods**). Systems were required to proceed autonomously from protocol interpretation to clinical evidence end to end. Generic and medical LLMs frequently failed before quantitative analysis because of poorly constructed ascertainment strategies, incomplete eligibility capture, incompatible data, and errors accumulated along the research trajectory, yielding valid clinical evidence in only 3.8% of evaluations for GPT-5 and 2.9% for Llama-3-70B (**Fig. 2e**). Applying the OpenEBM inference pipeline converted otherwise erroneous trajectories into methodologically valid research conduct, increasing end-to-end validity to 54.6% for GPT-5 and to 41.4% for Llama-3-70B. OpenEBM-8B produced valid evidence in 90.7% of evaluations across 33 medical and health domains.

#### Human performance on OpenEBM-Bench

Human experts remained a strong baseline across the full research trajectory (**Figs. 2a–d**). They generally outperformed OpenEBM-GPT-5 and OpenEBM-70B, whereas OpenEBM-8B matched human experts overall and exceeded them in maximum-sensitivity trial ascertainment and risk-of-bias assessment. In stages conventionally conducted through independent dual review followed by arbitration, OpenEBM showed higher agreement between independent review agents than paired human experts in first-pass triage, eligibility adjudication, arm-level data collection, and risk-of-bias and certainty-of-evidence assessment (**Fig. 3a**). In contrast-level data collection where decisions are more factual and allow less interpretive discretion, agreement was comparable between OpenEBM-8B and human experts.

**Fig. 3.**
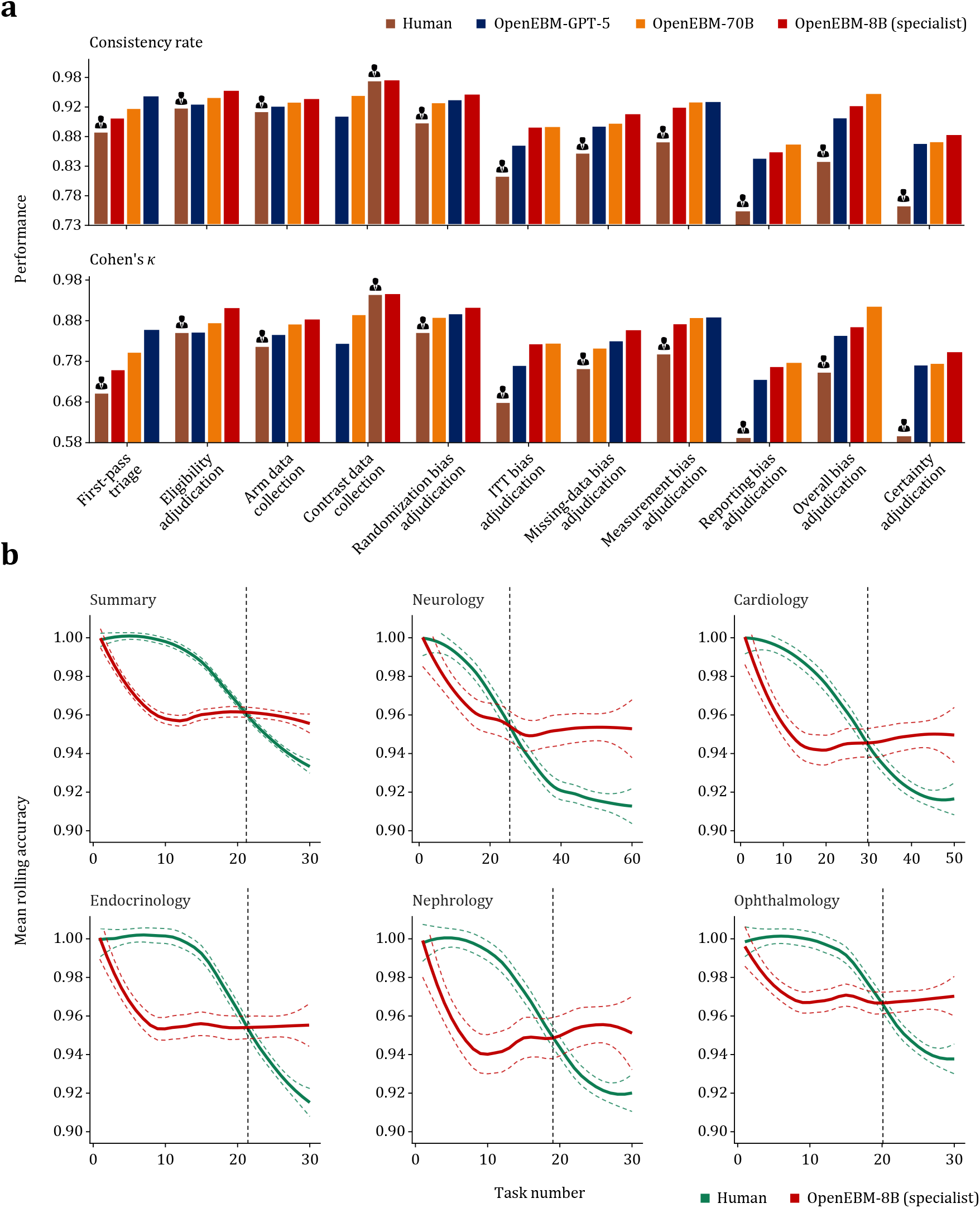
Human consistency and rolling accuracy across sequencing tasks. (**a**), Consistency across stages conventionally requiring duplicate review. OpenEBM and human experts are compared by consistency rate and Cohen’s **κ** before arbitration across first-pass triage, eligibility adjudication, data collection, and bias and certainty assessment. (**b**), Performance over sustained eligibility adjudication. Rolling accuracy trajectories show human experts and OpenEBM across sequential screening tasks in five clinical domains and in the aggregate summary. Dashed lines indicate 95% confidence intervals, and vertical dashed lines mark the point at which human accuracy declined below the level maintained by OpenEBM. OpenEBM shows stable performance across task volume, whereas human accuracy declines with sustained workload.

We observed evidence of cognitive fatigue in human experts. Trials were randomly permuted for each expert independently to eliminate ordering and difficulty confounding (**Methods**). During the initial phase of eligibility adjudication, human experts achieved near-perfect accuracy that exceeded OpenEBM. However, accuracy declined over the first 10 judgements to below the level OpenEBM maintained throughout (**Fig. 2b**). Most misclassifications in this phase were false positives, in which ineligible trials were incorrectly advanced to data collection, consistent with reduced discriminability under increasing cognitive load. OpenEBM showed no detectable performance drift across the sequence, holding consistent accuracy regardless of task volume. Its advantage over human experts thus arises partly from sustained consistency across the high-throughput, repetitive judgements the process demands.

OpenEBM also accelerated the research process. For 30 highly relevant trials, each human expert required an average of 4.1 hours to complete eligibility adjudication, or 16.4 expert-minutes per study under standard dual-review. By contrast, OpenEBM completed this stage in 4.8 minutes, or 9.5 seconds per trial, reducing the required time by 99.0%. Comparable gains held across other stages, where OpenEBM reduced human time by 43.0–99.9%.

### OpenEBM matches expert reasoning traces and clinical conclusions

Beyond automated benchmarking, we conducted blinded evaluations comparing OpenEBM outputs with those of multidisciplinary Cochrane expert teams, whose work sets the reference standard for clinical evidence generation. The evaluation spanned 50 held-out studies performed by OpenEBM across five domains and 19,500 fine-grained judgements, covering reasoning traces generated during the research trajectory and final evidence reports (**Methods**). One pool of independent evaluators scored matched, blinded output pairs for accuracy, completeness, reasoning quality, and conciseness, while a separate, non-overlapping evaluator pool performed a Turing-style test (**Methods**).

#### Expert quality evaluation

OpenEBM and Cochrane-authored outputs occupied a narrow quality range, with mean scores between 4.21 and 4.62 on a five-point scale. No pairwise difference exceeded 0.3 on any dimension, and no output category showed a dominant advantage for either source (**Table 1**). Within reasoning traces, OpenEBM tended to provide more complete justifications by exhaustively enumerating supporting evidence, whereas Cochrane expert teams foregrounded salient considerations more selectively. Neither strategy yielded a net quality advantage. Certainty-of-evidence reasoning showed the most balanced preference distribution (tie rate, 34.3%), consistent with the highly structured criteria governing this stage. Risk-of-bias reasoning showed the lowest tie rate yet a near-symmetric preference split, indicating that evaluators perceived distinct strategies of comparable quality.

When translating evidence into clinical conclusions, OpenEBM drew the strongest preference from independent evaluators. They favored OpenEBM over Cochrane expert teams in 44.2% versus 40.6% of comparisons, with advantage primarily attributed to completeness (+0.14) and reasoning quality (+0.11) despite lower conciseness (−0.12). The exhaustive reasoning that lengthened intermediate traces thus translated into more comprehensive conclusions at the most clinically consequential stage. Procedural narratives and evidence conclusions both showed lower tie rates than reasoning traces, suggesting that evaluators more readily perceived stylistic differences in longer outputs; however, these differences carried no consistent quality preference for either source.

#### Turing-style test

We next asked whether evaluators could distinguish OpenEBM outputs from those produced by multidisciplinary Cochrane expert teams. A separate pool of independent evaluators was shown individual reasoning traces and evidence reports without source labels and asked to judge whether each originated from OpenEBM or a Cochrane expert team, while rating their confidence (**Methods**). Evaluators were informed that either source was possible but were not given the source distribution.

Source-identification accuracy ranged from 48.4–54.8% across all categories, indistinguishable from chance (**Table 2**). Reasoning traces were the hardest to attribute, with three of four categories falling below chance, consistent with the tighter evidential and methodological constraints on these stages that limit divergence. Evidence reports were marginally more identifiable, yet the most distinguishable category remained within five percentage points of chance. Evaluator confidence was poorly calibrated to accuracy. The clearest dissociation appeared in clinical conclusions, where evaluators expressed greater confidence when they were wrong than right. These findings show that OpenEBM produced reasoning traces and clinical conclusions matching Cochrane expert teams in quality and indistinguishable from them under blinded attribution.

### OpenEBM reproduces, updates, and generates *de novo* clinical evidence

To evaluate whether OpenEBM can generate clinically consequential evidence in real-world practice, we applied the system in three progressively demanding settings: reproducing established clinical evidence, updating it as evidence bases evolved, and generating *de novo* evidence where none existed. After confirming fidelity in reproduction and update across diverse domains, we applied OpenEBM to an unaddressed question in current guidelines: the efficacy and safety of neoadjuvant chemotherapy for locally advanced rectal cancer (**Methods**).

#### Reproduction and update of established clinical evidence

We first assessed whether OpenEBM could reproduce established clinical evidence by applying it to three recently published studies, across ophthalmology, psychiatry, and cardiology (**Methods**). These included studies of fenofibrate for diabetic retinopathy,^15^ omega-3 fatty acid supplementation for adolescent depression^16^, and preoperative statin therapy for cardiac surgery^17^, which were completed an average of 38.8 months after protocol release. For each study, OpenEBM received the protocol and autonomously conducted the full research life cycle, without access to any intermediate decisions or final outputs. Literature search dates were matched to those reported in the corresponding published studies.

Across all three studies, OpenEBM identified every eligible trial included by the human expert teams. During data collection, OpenEBM autonomously navigated complex methodological decisions that typically require expert judgement, including unit-of-analysis issues, recovery of missing statistics, and reconstruction of intention-to-treat denominators after postrandomization attrition. Of 586 datapoints collected, 530 (90.4%) matched the human expert teams’ decisions exactly or within digit rounding (**Supplementary Table S2**). Post hoc review attributed the remaining 48 discrepancies to either defensible methodological differences or likely oversights in the published studies. Two arose when OpenEBM recovered means from medians using established Wan’s method, whereas 46 reflected OpenEBM’s handling of postrandomization attrition when outcome assessment supported intention-to-treat reconstruction. After accounting for these differences, OpenEBM achieved near-perfect concordance with the published studies.

These intermediate discrepancies did not alter the clinical conclusions. OpenEBM reproduced the published studies with near-complete concordance across 54 prespecified primary estimates and 65 subgroup analyses, preserving direction and magnitude in all cases (**Fig. 4a** and **Supplementary Table S3**). Among the 54 primary estimates, 50 (92.3%) were reproduced within 5% relative differences of the published estimates. Most larger relative deviations arose for very small, non-significant effects, for which absolute differences remained minimal. One primary estimate showed a larger deviation because the human expert team had incorporated unpublished data obtained through author contact, which lay outside the scope of OpenEBM. The same source accounted for most residual differences in heterogeneity and subgroup estimates, typically increasing within-group heterogeneity in the published study through the inclusion of otherwise unavailable data (**Fig. 4b** and **Supplementary Table S4**). All interaction estimates reflecting between-subgroup differences were reproduced concordantly.

**Fig. 4.**
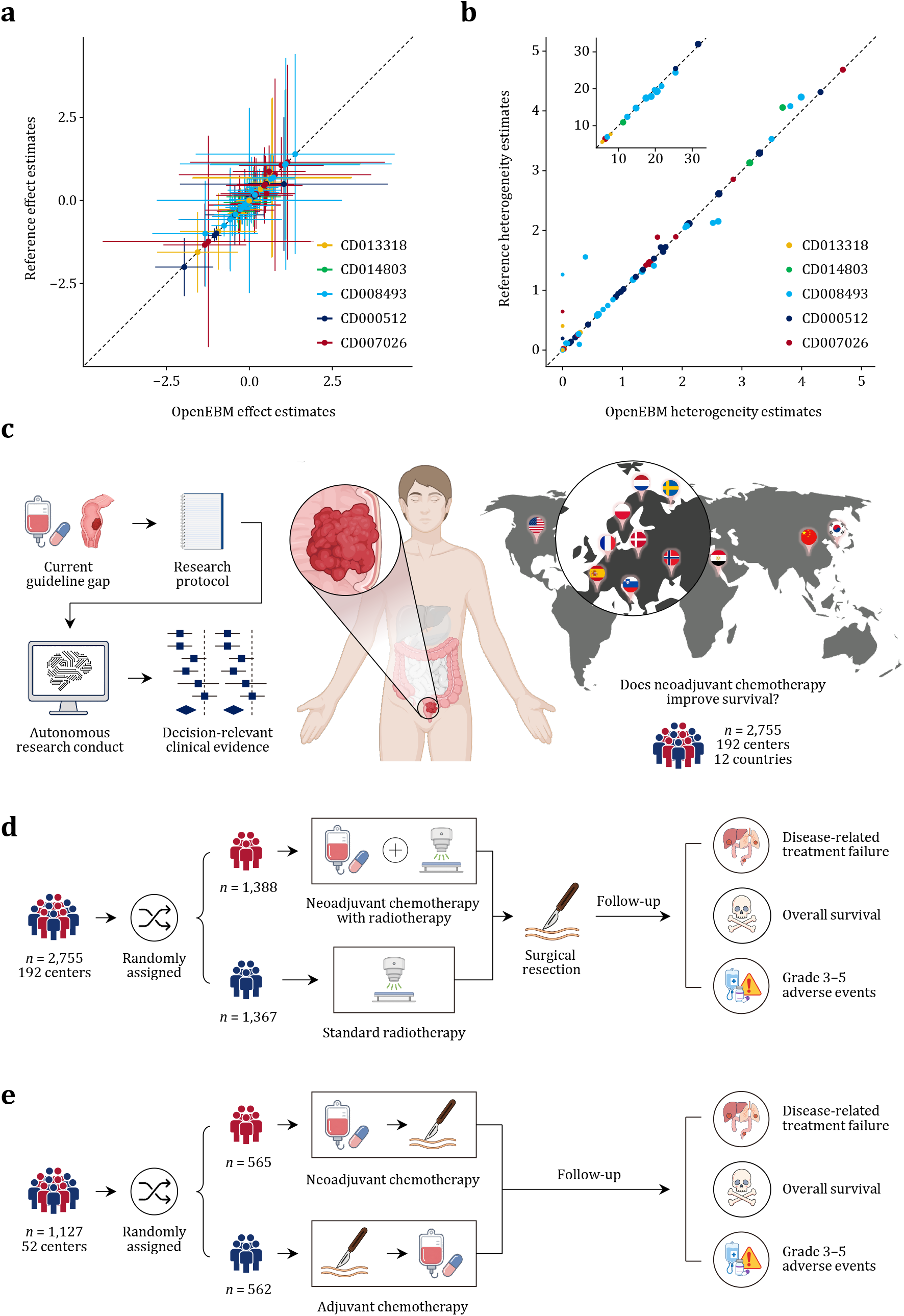
Reproduction, update and *de novo* generation of clinical evidence. (**a**), Concordance of effect estimates between OpenEBM and the reference studies across reproduction and update experiments. Points denote paired effect estimates, error bars denote uncertainty intervals, and the dashed line indicates perfect agreement. (**b**), Concordance of heterogeneity estimates between OpenEBM and the reference studies. The inset expands the range of larger heterogeneity estimates; circle diameters denote degrees of freedom, and the dashed line indicates perfect agreement. (**c**), *De novo* clinical evidence generation for locally advanced rectal cancer. Starting from a guideline evidence gap and research protocol, OpenEBM autonomously conducts the full research life cycle and generates decision-grade clinical evidence from randomized trials conducted across 192 centers in 12 countries. (**d**), Randomized trials comparing neoadjuvant chemotherapy plus radiotherapy against standard radiotherapy, followed by surgical resection and follow-up for disease-related treatment failure, overall survival, and grade 3–5 adverse events. (**e**), Randomized trials comparing preoperative neoadjuvant chemotherapy with postoperative adjuvant chemotherapy, with follow-up for disease-related treatment failure, overall survival, and grade 3–5 adverse events.

We then evaluated whether OpenEBM could update established evidence when evidence bases evolved. For two published studies with published update histories, OpenEBM independently interpreted the amended protocol and conducted the updated research (**Methods**). In the cerebrolysin study for acute ischemic stroke,^18^ OpenEBM identified protocol amendments that expanded the intervention scope to include cortexin and added neuroimaging confirmation of stroke as an eligibility requirement, leading to the inclusion of one additional trial and exclusion of another relative to the earlier study. In the study of hemoglobin concentration thresholds for transfusion in very low-birthweight infants,^19^ OpenEBM identified two newly eligible trials whose incorporation increased the evidence base and raised certainty in the resulting conclusions. OpenEBM also flagged a potential oversight in the published update, where intention-to-treat denominator reconstruction for retinopathy of prematurity was not supported by the reported data, although correcting this issue did not change the study conclusion (**Figs. 4a–b**). Across both studies, OpenEBM reproduced all material conclusion changes reported in the published updates.

#### De novo generation of clinical evidence

We next tested whether OpenEBM could complete a *de novo* study for a clinically active question. We applied the system to a registered protocol^20^ on neoadjuvant chemotherapy for locally advanced rectal cancer, for which no such evidence had yet been generated (**Fig. 4c**). Although current clinical guidelines recommend this treatment based on surrogate pathological endpoints, its direct survival benefit, optimal timing, and safety profile remain unresolved.^12–14^ Drawing on evidence from seven randomized trials involving 2,755 patients across 192 centers in 12 countries, OpenEBM produced clinical evidence that clarified the benefits, harms, and broader treatment strategy of neoadjuvant chemotherapy for locally advanced rectal cancer (**Supplementary Tables S5–S11**). The full study was completed in 115.3 minutes. A multidisciplinary validation panel independently reviewed the methodological conduct and clinical interpretation of all stages of the study (**Methods**).

Compared with no neoadjuvant chemotherapy (**Fig. 4d**), neoadjuvant chemotherapy improved overall survival (HR = 0.76; 95% CI: 0.64 to 0.91; four trials, 2,487 patients; high-certainty evidence) and reduced disease-related treatment failure (hazard ratio [HR] = 0.75; 95% CI: 0.60 to 0.94; one trial, 912 patients; high-certainty evidence) over three years. There was no evidence that this survival benefit differed according to whether chemotherapy was delivered before or after radiotherapy or whether it was combined with short-course radiotherapy or chemoradiotherapy (**Supplementary Table S9**). OpenEBM also identified a safety signal not reflected in current clinical guidelines, showing that neoadjuvant chemotherapy was accompanied by increased serious adverse events (odds ratio [OR] = 1.59; 95% CI: 1.16 to 2.18; seven trials, 2,734 patients; moderate-certainty evidence).

Having established a survival advantage for neoadjuvant chemotherapy, OpenEBM next examined whether delivering the same systemic treatment before surgery is superior to postoperative adjuvant chemotherapy, which represents the principal alternative strategy in current practice (**Fig. 4e**). In direct comparisons of neoadjuvant versus adjuvant chemotherapy, preoperative delivery improved overall survival (HR = 0.66; 95% CI: 0.49 to 0.89; two trials, 1,060 patients; high-certainty evidence) over three years, consistent with a benefit of earlier systemic exposure that improves treatment completion and controls micrometastatic disease before postoperative recovery compromises chemotherapy delivery.^21, 22^ As in the first comparison, this survival gain was accompanied by a higher risk of severe adverse events (OR = 1.91; 95% CI: 1.19 to 3.04; three trials, 1,119 patients; moderate-certainty evidence).

These findings resolve two central questions current guidelines leave unsettled: whether neoadjuvant chemotherapy confers a survival benefit, and whether preoperative delivery outperforms postoperative. They therefore identify preoperative delivery as the preferred systemic strategy for locally advanced rectal cancer while exposing the toxicity cost that should govern its selective use. By supplying the evidentiary foundation that guidelines lack, OpenEBM shows that autonomous evidence generation can refine clinical recommendations at the frontier of unresolved practice.

## Discussion

We show that decision-grade clinical evidence can be generated autonomously with expert-level quality and reliability. We introduced OpenEBM-Bench, a multidisciplinary benchmark evaluating autonomous clinical evidence generation across the full research life cycle, on which OpenEBM achieved a 90.7% end-to-end success rate in generating valid clinical evidence, whereas state-of-the-art models rarely succeeded. OpenEBM matched or exceeded human experts across most stages, and domain experts identified the source of its reasoning at rates indistinguishable from chance. Across five clinical domains, the system reproduced completed studies from multidisciplinary human expert teams with 92.3% concordance of primary estimates and generated *de novo* evidence that neoadjuvant chemotherapy improves overall survival in locally advanced rectal cancer, a question left unresolved by current guidelines.^12–14^ Research that has traditionally required years of effort was completed autonomously in less than two hours.

The collapse in time and cost begins to realize a promise as old as EBM itself. When David Sackett wheeled his evidence cart through hospital wards three decades ago, carrying a laptop and printer to bring appraised evidence to the bedside, he embodied the founding aspiration that every clinical decision should rest on evidence.^23, 24^ That aspiration has remained only partially fulfilled because the evidence itself could not be produced quickly enough to keep pace with clinical need. However, when evidence can instead be generated on demand, it becomes a continuously renewable resource rather than a decaying snapshot, as our updating experiments show by regenerating current evidence directly from evolved evidence bases. This shift also lowers the barrier that has left urgent clinical questions without decision-grade evidence, spanning both the neglected margins of medical practice—rare diseases, under-resourced settings, and under-represented populations—and contested questions at the center of mainstream practice. Our *de novo* study illustrates this potential by resolving a survival question left unresolved by current guidelines, while surfacing a serious-adverse-event signal those guidelines do not reflect.^12^

Earlier and contemporaneous efforts underscore the difficulty of this goal. One line of work automates selected stages of the process, yet contemporaneous benchmarking shows that these dependent stages do not hold up when chained.^25–28^ Even the direction of the final estimate, a three-way judgement far short of a quantified and graded result, is reached by the strongest configuration only 61.4% of the time and by most near chance.^28^ A second line of work reproduces the guideline-development workflow with humans in the loop, but aligns outputs to existing recommendations rather than generating clinical evidence directly.^29^ Contemporaneous work alongside ours assembles proprietary LLMs to reproduce Cochrane studies, but still relies on substantial human correction and covers a restricted span of the research trajectory, leaving trial ascertainment, certainty-of-evidence assessment, and clinical conclusion beyond its reach.^30^ None conducts the research itself. OpenEBM does, proceeding autonomously across the full research life cycle, bolstered by a specialist model trained on expert EBM research trajectories, and producing *de novo* clinical evidence where none previously existed.

These findings have implications beyond medicine for broader knowledge production where validity depends on explicit methodological constraints and expert-practice supervision. Within this framework, the same model backbone that failed in unconstrained use succeeded once embedded in a protocol-grounded inference pipeline, and a compact specialist model surpassed much larger generic models. Scale alone falls short because these tasks reward methodological defensibility rather than the surface plausibility at which larger models excel. The same paradigm extends to other disciplines confronting the most pressing global challenges, including climate change, energy transitions, biodiversity loss, antimicrobial resistance, and poverty eradication.^3, 4^ While the questions to be answered span from climate interventions to ecosystem consequences, the underlying research structure requiring faithful adherence to an explicit protocol-grounded framework is shared. Wherever valid knowledge production depends on such constraints, encoding methodology and learning from expert practice may provide a transferable paradigm for autonomous research. Given a discipline’s foundations, such a system could be expected to generate pressing evidence within that field as OpenEBM within ours.

These capabilities carry responsibilities that warrant explicit consideration. Although OpenEBM matches human expert teams, the accessibility that could accelerate the closing of genuine evidence gaps could equally enable proliferation of flawed evidence without oversight. OpenEBM mitigates this risk through auditability, where each research action is constrained by explicit rules and traceable to its evidentiary basis, allowing human review of both the decision and the evidence supporting it. Its current scope also defines a clear path for further development. First, although our specialist model was trained on a Llama backbone, the OpenEBM framework is model-agnostic and further gains are likely from combining stronger reasoning backbones with EBM research-trajectory pretraining and protocol-grounded inference. Second, its present reach is constrained by the openness of the scientific literature, as many trial reports remain behind paywalls; such reports can already be incorporated when supplied directly, and continued progress in open science should reduce this constraint. Finally, OpenEBM is currently grounded in randomized-trial evidence, and extension to network and non-randomized designs remains to be established.

In summary, OpenEBM shows that autonomous systems can generate decision-grade clinical evidence that matches expert quality. By transforming evidence synthesis from a recurrent bottleneck into a continuously executable research capability, and by establishing a paradigm of protocol-grounded generation in the research trajectories of expert practice, OpenEBM points toward a future in which the rigorous knowledge production that underpins medicine, and evidence-based decision-making more broadly, can be carried out at the scale and continuity that real-world practice demands.

## Methods

### Overview of OpenEBM

We first provide an overview of the evaluation framework that consists of four complementary components. First, automated benchmark evaluation was conducted across 33 medical and health domains, using reference annotations derived from 200 domain-stratified evidence-synthesis studies, enabling systematic assessment of both methodological decision-making and full research life cycle execution. Second, blinded human expert evaluation was performed on 50 matched studies conducted by OpenEBM and multidisciplinary Cochrane expert teams, where independent domain-matched experts assessed research conduct across trajectories and reporting quality. Third, a Turing-style source-identification test was performed using the same outputs, where another pool of expert panel judged whether each research conduct originated from OpenEBM or Cochrane expert teams. Fourth, external validity was assessed through reproduction, updating, and *de novo* experiment, each described in detail below.

Evidence synthesis research differs fundamentally from the question-answering and narrative-summarization tasks at which large language models excel. Rather than retrieving or compressing existing knowledge, it requires execution of a complete research process in which protocol-bound decisions are made sequentially across mutually dependent stages, progressively transforming a body of primary trials into decision-grade clinical evidence that exists in no individual source. OpenEBM is an autonomous system designed for this setting, executing the full research life cycle as a protocol-grounded sequential decision process.

We formalize an evidence-synthesis study as a research trajectory driven by a protocol *π*, which specifies the research question, objective, and other study-defining constraints. Given *π*, OpenEBM produces a finite trajectory of research actions:

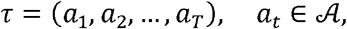

where *τ* is the full research trajectory and each action *a*_*t*_ is a research decision drawn from the space *A* of admissible actions. This action space spans protocol interpretation, trial ascertainment, eligibility adjudication, data collection, risk-of-bias assessment, quantitative evidence generation (including primary analyses and exploratory analyses where applicable), certainty-of-evidence assessment, and clinical conclusion generation. To represent the cumulative and path-dependent nature of this process, we associate each step with a research state *s*_*t*_, which stores all intermediate products available before action *a*_*t*_ is taken. The initial state is constructed directly from the protocol as *s*_0_ = *Init*(*π*). The state then evolves recursively as the study proceeds:

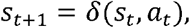

where *δ* is a transition operator that updates the current research state *s*_*t*_ with the newly produced action *a*_*t*_. Operationally, *δ* incorporates newly identified trials, eligibility decisions, collected datapoints, risk-of-bias assessments, quantitative evidence outputs, and certainty judgements into the evolving body of evidence.

OpenEBM generates each research action using model *p*_*θ*_ with parameters *θ*, conditioned on the protocol, the current research state, and a step-specific constraint set that encodes the methodological rules governing the action:

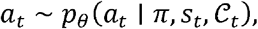

where *C*_*t*_ is the constraint set active at step *t*, confining each generation to protocol-consistent, methodologically defensible actions. Its derivation from the protocol and its role in the inference pipeline are described below. For selected stages, OpenEBM additionally performs independent dual generation followed by arbitration, mirroring the duplicate review procedures commonly used in expert-conducted practice, as described in the same section.

The complete process expresses the research as a protocol-grounded, autoregressive execution of the research trajectory:

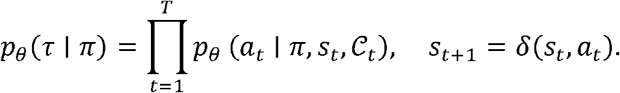

The clinical conclusion is read out from the terminal research state. We denote this output by ℰ = *Readout*(*s*_*T*+1_), which includes the clinically relevant evidence products generated by the completed study, including effect estimates, heterogeneity estimates, uncertainty estimates, risk-of-bias and certainty-of-evidence assessment, and their associated interpretation outputs. OpenEBM realizes this framework through three components: a domain-specialized corpus that provides expert practice of EBM research trajectories, a specialist model trained on those trajectories, and an inference pipeline that enforces protocol-grounded generation, each described in turn below.

### OpenEBM-Corpus

Training a specialist model to conduct evidence-synthesis research and generate decision-grade clinical evidence requires supervision at the level of research actions. A model exposed only to completed reports can learn to imitate their language rather than the sequence of protocol-grounded methodological judgements through which clinical evidence is produced. OpenEBM-Corpus is constructed to provide this supervision directly by representing evidence-synthesis studies as annotated EBM research trajectories, in which each reference action is paired with the research state and methodological constraints under which it was taken.

We compiled the corpus based on Cochrane studies published since 2000. We selected Cochrane because it constitutes the most methodologically standardized body of evidence-synthesis practice across medical and health domains. Comparative studies have found Cochrane studies to be methodologically more rigorous than non-Cochrane studies published in other journals.^31–35^ More importantly for trajectory supervision, Cochrane studies are produced under an explicitly codified methodological framework defined by the Methodological Expectations of Cochrane Intervention Reviews (MECIR), which govern the research life cycle from protocol to initial evidence and subsequent updating. For trajectory supervision, the Cochrane therefore provides a uniquely standardized source that we systematically reconstruct into annotated EBM research trajectories under explicit methodological governance.

From each Cochrane study, we reconstruct a full EBM research trajectory by reverse-parsing the published studies into the ordered sequence of research actions that generated it. These actions include trial ascertainment strategies that define the candidate evidence base, eligibility adjudication decisions together with their recorded reasoning traces, collected trial characteristics and outcome data, risk-of-bias assessments and supporting reasoning traces, specifications for quantitative evidence generation, and certainty-of-evidence assessments with their downgrading rationales. For recent Cochrane studies, much of this structure is available in machine-readable form and can therefore be parsed directly. For earlier studies that predate structured data deposition, we recover the same action sequence from the published studies and accompanying materials through a dedicated extraction pipeline. Each recovered action is then normalized into a structured representation.

Although each Cochrane study defines a complete research trajectory, OpenEBM-Corpus is organized at the level of decision instances, pooling corresponding steps across studies so that each research action type is represented by a large, supervised set. Formally, the corpus is represented as

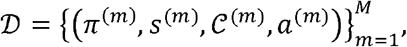

where *D* is the full corpus, *M* is the total number of annotated decision instances, *π*^(*m*)^ is the protocol associated with the instance, *s*^(*m*)^ is the research state in which that action was taken, *C*^(*m*)^ is the methodological constraint set, and *a*^(*m*)^ is the expert research action. This representation converts each published evidence-synthesis study from the Cochrane library into a collection of supervised research actions, each grounded in its protocol, state and methodological constraints. These step-level instances provide the direct supervision used for the specialist model described in the following section. In total, OpenEBM-Corpus includes 5.7 million annotated expert decision steps spanning 33 medical and health domains.

### EBM research-trajectory pretraining

OpenEBM-Corpus enables the conversion of research into a supervised learning problem in which the model learns to predict the expert research action at each step from the protocol, the current research state, and the governing methodological constraints. We train OpenEBM-8B by fine-tuning a Llama-3-8B backbone on these annotated EBM research trajectories. Each trajectory is decomposed into its constituent decision instances, such that every training example presents the task-relevant context, including the research objective, the active methodological constraints, and the pertinent excerpt of the primary trial report, paired with the expert action as the supervision target.

Training is performed with a conditional generation objective. For a study trajectory *i*, let 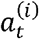 denote the expert action at step *t*, let 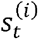 denote the corresponding research state, let 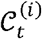 denote the step-specific methodological constraint set, and let *π*^(*i*)^ denote the protocol. The training objective minimizes the negative log-likelihood of expert actions across the corpus:

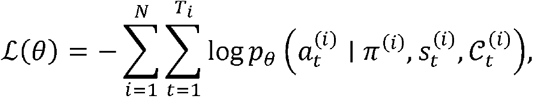

where the target action 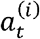 is the structured output at that stage, including trial ascertainment strategies, eligibility adjudication decisions and rationales, characteristic profiles, data field collection, risk-of-bias assessments and rationales, quantitative effect and heterogeneity estimates, certainty-of-evidence assessments and rationales, or clinical conclusions depending on the step. Conditioning every step on the explicit constraint set 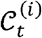 during training aligns pretraining with protocol-grounded generation, such that the same conditioning mechanism can enforce methodological compliance during inference. Restricting the input context to the task-relevant excerpt of the trial report further concentrates supervision on the evidence pertinent to the action being learned.

To prevent leakage and enable temporally prospective evaluation, the training corpus was capped at 1 June 2023. All downstream evaluation studies were published after this cutoff and were excluded in their entirety, including their registered protocols, earlier published versions, linked trial reports, and any step-level intermediate supervision derived from their reconstructed research trajectories. Because leakage can occur through shared primary trials, any linked trial reports recorded as included or excluded in these temporally held-out studies were identified using trial identifiers, digital object identifiers, and report titles. Any reconstructed trajectories linked to these trials or reports were then excluded from corpus supervision.

Since our specialist model was initialized from a Llama-3-8B backbone whose disclosed pretraining cutoff was March 2023, preceding both the predefined corpus supervision cutoff and all post-cutoff evaluation studies, it is unlikely to have been exposed during pretraining to any downstream evaluation studies published after June 2023. However, pretraining exposure cannot be completely audited for other third-party foundation models used as comparators, particularly proprietary models whose training corpora are not fully disclosed. We therefore interpret cross-model comparisons conservatively, as any residual exposure of comparator models to post-cutoff evaluation studies or linked trial reports would be expected to inflate their performance. In contrast, OpenEBM-8B was trained exclusively on temporally capped and study-family-excluded data, ensuring by construction that it had no exposure to any evaluation studies or associated supervision. This guarantees that any leakage bias acts against, rather than in favor of, the observed performance advantage of our model.

We adopted parameter-efficient fine-tuning with low-rank adaptation, updating a compact set of adapters while preserving the broad capabilities of the base model. Adapters of rank 32 with scaling factor of 8 and dropout of 0.05 were applied to 32 layers. Training was performed for up to two epochs over 5.7 million serialized step-level training instances derived from OpenEBM-Corpus trajectories, with a learning rate of 5×10^−5^ under a cosine learning-rate schedule with warmup and early stopping on a held-out split. Each action was serialized as a structured JSON object, and no new special tokens were introduced into the model vocabulary; field delimiters were represented as ordinary strings that the model learned to produce reliably during training.

### OpenEBM inference pipeline and protocol-grounded generation

Given a protocol, the OpenEBM inference pipeline resolves the governing methodological rules, advances through the research trajectory by supplying each step with its appropriate context and constraints, invokes the specialist model for methodological judgements, delegates all quantitative operations to verified procedures, applies independent duplicate review where reliability requires it, and reads out the resulting clinical evidence from the terminal research state.

Protocol-grounded generation begins by resolving the protocol into an explicit rule system. The pipeline maintains a structured methodological plan in which each research stage carries a default specification encoding standard evidence-synthesis practice. These defaults include, for example, bibliographic sources to be queried, handling of unit-of-analysis issues, admissible effect measures for a given outcome type, and synthesis models. A protocol-resolution operator *ρ* then parses the study protocol *π* and overrides these defaults with the protocol-specific decisions it contains. Optional researcher-supplied directives may in turn override protocol-derived defaults. This produces a fully resolved rule set ℛ = *ρ*(*π*). At each step of the research trajectory, the instantiation operator κ selects from ℛ the subset of rules applicable to the current research stage and state, producing the local constraint set *C*_*t*_ = κ(ℛ,*s*_*t*_) that governs the decision.

These constraints act on generation in two complementary ways. For actions whose outputs must belong to a fixed set of admissible options, *C*_*t*_ restricts the global action space *A* to a feasible subset 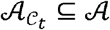. These cases include, for example, admissible effect measures dictated by outcome type, classifications of trial design, and other decisions with explicitly bounded option sets. The pipeline enforces such constraints by validating the model’s structured output against 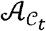 and rejecting generations that are not methodologically compliant. For actions that instead require contextual methodological judgement, such as grounding an exclusion decision or risk-of-bias decision in the content of a trial report, *C*_*t*_ enters the conditioning context as structured directives that steer the action within the admissible space.

A defining property of the pipeline is that the model performs methodological judgement, whereas all quantitative operations are delegated to deterministic and verifiable procedures. While the model determines eligibility and risk-of-bias judgements; it does not generate numerical estimates directly. Steps including combination or splitting of arms in multi-arm trials, recovery of missing statistics through available data, quantitative synthesis under specified models are all executed by verified computational routines acting on the model’s structured outputs. This separation confines the model to the methodological reasoning for which it is trained and removes numerical estimation from the space of free-form generation.

For research stages in which standard evidence-synthesis practice requires independent verification, including eligibility adjudication, data extraction, and risk-of-bias and certainty-of-evidence assessment, the pipeline performs duplicate generation followed by arbitration. Two candidate actions are sampled independently from the same conditional distribution at temperature 0.7:

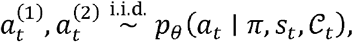

where 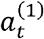 and 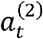 are the two independently generated candidate actions for step *t*. If the two candidates agree, the action is accepted directly. If they disagree, the pipeline invokes an arbitration operator *V*, which re-evaluates the competing candidates under the same protocol, state and constraint context at a lower temperature of 0.2, favoring a more stable resolution:

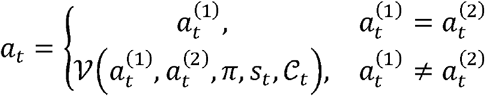

This dual-agent arbitration procedure mirrors the independent duplicate review commonly used in expert-conducted research and contributes to the high internal consistency of OpenEBM at these stages.

### OpenEBM-Bench

Existing evaluations of evidence synthesis typically target isolated subtasks within narrow clinical domains, failing to measure whether a system can maintain methodological competence across dependent stages and various domains. OpenEBM-Bench was designed to address these gaps by reconstructing the full research life cycle from 200 temporally held-out and domain-stratified studies spanning 33 medical and health domains and evaluating systems along the actual causal chain of research conduct through 24 action checkpoints and 15 chained or end-to-end trajectories, yielding 122,800 evaluations per system. The benchmark supports comparisons between state-of-the-art proprietary and open models, with and without the OpenEBM inference pipeline, thereby isolating the contributions of model scale, protocol-grounded inference, and EBM research-trajectory pretraining. To ensure comparability, all systems received identical inputs at each research action, and generic LLMs were prompted to return the same structured outputs without access to the OpenEBM inference pipeline.

OpenEBM-Bench is organized into three evaluation regimes of increasing inter-stage dependency. In the stepwise regime, each research action is evaluated in isolation: gold-standard outputs from all preceding stages are supplied as input, and the system generates only the action under test. This regime measures the intrinsic competence of each stage independently of upstream error and provides the reference against which later error propagation is quantified. In the chained regime, the benchmark evaluates three critical interfaces—trial ascertainment to eligibility adjudication, eligibility adjudication to data collection, and data collection to quantitative evidence generation—by carrying forward the system’s own upstream outputs rather than gold-standard inputs. The resulting performance drop relative to the stepwise regime quantifies the accumulation of error across dependent research stages. In the end-to-end regime, all intermediate gold standards are withheld, requiring the system to proceed autonomously from protocol interpretation to final clinical evidence generation.

All benchmark evaluations were run five times and averaged to account for stochasticity in model generation. Evaluation metrics were matched to the type of research action being assessed (**Supplementary Table S1**). Structured classification outputs, including atomic search terms and eligibility adjudication decisions, were evaluated using recall, precision, and F1 score. Ordinal judgements, including risk-of-bias and certainty-of-evidence assessments, were evaluated using accuracy, macro-F1 and quadratic weighted κ. Quantitative outputs, including effect sizes, standard errors, and heterogeneity statistics, were evaluated by within-tolerance accuracy and mean absolute error. Concordance before arbitration was assessed by raw agreement rate and Cohen’s κ between independently generated candidate actions. For trial ascertainment, evaluation of search-strategy construction was confined to the three bibliographic databases mandated by the MECIR. Reference retrieval sets were constructed by running the reported search strategies up to the reported search dates.

Free-text reasoning traces, including exclusion rationales and the justifications accompanying risk-of-bias and certainty-of-evidence assessments, were additionally evaluated using an automated LLM judge against four criteria: whether the rationale cited relevant information from the source trial; whether it explained how the information supported the judgement; whether it was logically consistent with the gold-standard judgement; and whether it contained no irrelevant or fabricated content. This analysis served as an auxiliary validation of reasoning consistency and was not used for any primary performance comparisons. For characteristics of included studies, evaluation of supporting explanations was not required. To assess the reliability of this judge, we used a frontier model (DeepSeek-V4-Pro; not included among evaluated systems) as the automated evaluator, and two domain experts independently rated a random sample of 200 reasoning traces against the same criteria. The two experts agreed with one another at an average Cohen’s κ of 0.77 across tested stages, whereas the automated judge agreed with the human consensus at an average Cohen’s κ of 0.73. Formal evaluation of traces was conducted in the blinded human expert evaluation described below.

The 200 studies used for trajectory benchmarking were selected by proportional stratified sampling across the 33 medical and health domains represented in the Cochrane database. The number of studies drawn from each domain was set in proportion to that domain’s prevalence in the database, subject to a minimum of two studies per domain and a maximum of 20 studies for the largest domain (**Supplementary Table S12**). This design preserved coverage across the full breadth of medical and health domains while preventing any single domain from dominating the benchmark. The allergy and immunology domain was the only exception where only one eligible study was available after the training cut-off.

### Human performance and evaluation

We recruited 75 domain experts with at least five years of research experience and at least three peer-reviewed publications as first or last authors in the relevant field, spanning neurology, cardiology, nephrology, endocrinology and ophthalmology. Participants were divided into three non-overlapping panels of 25 experts, with five experts per domain assigned respectively to human performance on OpenEBM-Bench, quality evaluation, and the Turing-style test. The panels were kept separate so that task exposure in one evaluation could not influence performance in another. Across evaluations, the reference comparator was the corresponding Cochrane study, whose outputs were restructured into the same representation as the OpenEBM outputs, so that each comparison contrasted an autonomous study and an expert-conducted study for the same research question. All participants provided informed consent, and no personally identifiable information was collected.

For human performance assessment, recruited domain experts performed benchmarked research actions under the same stepwise evaluation conditions as the autonomous systems, with gold-standard outputs from all preceding stages provided as input. The five experts in each domain independently assessed all items. For stages conventionally requiring independent duplicate review, first-pass judgements were recorded before any discussion, and inter-rater agreement was quantified using Cohen’s κ across the ten pairwise combinations of the five domain experts. Discordant items were resolved by a domain-matched senior arbiter who was not involved in the first-pass assessments. Human performance was scored against the same gold standards and using the same metrics applied to the systems. For each expert, trials were independently randomly permuted to eliminate ordering effects. This enabled isolation of within-expert temporal effects from between-trial variability. Sequential performance was quantified by indexing decisions along the screening order and aggregating accuracy over sliding windows.

For quality evaluation, outputs were compared at the level of category-specific units. For each study, all outputs belonging to a given category were assembled into a single unit, and the OpenEBM and Cochrane outputs of that unit formed one matched comparison. These paired units were presented side by side, with source labels removed and presentation order randomized. The evaluated categories comprised reasoning traces generated during the research trajectory—study characterization, exclusion rationales, and risk-of-bias and certainty-of-evidence supports—as well as evidence reports, including clinical conclusions and procedural narratives. Each paired comparison was scored on four dimensions—accuracy, completeness, reasoning quality, and conciseness— using a five-point scale, together with an overall source preference or a tie. Each comparison was rated independently by the five experts in the corresponding domain. Average inter-rater agreement was Cohen’s 0.70 across raters and domains. Across all studies, categories, and scoring dimensions, this evaluation yielded 16,500 fine-grained judgements.

For the Turing-style test, the same category-level units were presented individually, without source labels, to a separate panel of five experts per domain. For any given study and output category, the OpenEBM and Cochrane outputs were never shown to the same evaluator; each evaluator saw only one version, ensuring that source attribution was based on an isolated output rather than on direct comparison between pairs. Domain experts were informed that each unit could originate either from OpenEBM or from a multidisciplinary Cochrane expert team but were not told the source distribution. They independently judged the source of each unit and rated their confidence on a five-point scale.

This evaluation yielded 3,000 source-identification and confidence judgements. For each output category, we computed source-identification accuracy and tested it against the 50% chance level using a two-sided binomial test.

### Reproduction, updating, and *de novo* evidence-synthesis studies

We evaluated whether OpenEBM can operate autonomously across the full research life cycle in three settings: reproducing established clinical evidence, updating published evidence as the evidence base evolved, and generating evidence *de novo*. For reproduction, we applied OpenEBM to three studies published after the training cutoff: fenofibrate for diabetic retinopathy (CD013318), omega-3 fatty acid supplementation for depression in children and adolescents (CD014803), and preoperative statin therapy for adults undergoing cardiac surgery (CD008493).^15–17^ For updating, we applied OpenEBM to two studies with published version histories whose latest updates were made after the training cutoff: hemoglobin concentration thresholds for transfusion in very low birthweight infants (CD000512) and cerebrolysin for acute ischemic stroke (CD007026).^18, 19^

In both reproduction and update settings, OpenEBM received only the registered protocol or the Methods section of the published study as the protocol proxy when no protocol was available. Literature search dates were matched to those of the corresponding study so that the evidence base was held constant. Trial ascertainment was restricted to MEDLINE, Embase, and CENTRAL; although some studies additionally searched other sources, these three databases were sufficient to recover all included trials. Any datapoints obtainable only through correspondence and not available from the published trial report or publicly accessible supplementary materials were treated as unavailable to OpenEBM. Concordance with the published study was assessed at the levels of eligible trials, collected datapoints, pooled effect estimates, heterogeneity estimates, and subgroup estimates. Discordant datapoints were reviewed post hoc to trace the source of each discrepancy.

For *de novo* evidence synthesis, we applied OpenEBM to a registered protocol on neoadjuvant chemotherapy for locally advanced rectal cancer (CD015231), which had passed rigorous peer review but for which no completed study yet existed.^20^ Starting from the protocol alone, OpenEBM autonomously constructed trial ascertainment strategies and retrieved 2,881 records, of which 237 reports were advanced after first-pass triage for full-text eligibility adjudication (**Supplementary Tables S5–S6**). This process identified 25 reports from 17 eligible trials; because the protocol specified that missing outcome was not an eligibility criterion,^20^ eight reports from seven trials contained extractable data. OpenEBM then performed data collection, risk-of-bias assessment, primary and subgroup analyses, heterogeneity analyses, certainty-of-evidence assessment, and produced the clinical conclusion (**Supplementary Tables S7–S11**).

A multidisciplinary validation panel comprising two colorectal oncologists, one colorectal surgeon, and two evidence-synthesis methodologists reviewed the *de novo* study using a structured validation form. Panel members independently assessed trial eligibility, data extraction, risk-of-bias assessment, statistical analyses, certainty-of-evidence assessment, and clinical interpretation. All panel members judged the primary survival conclusion to be clinically and methodologically defensible. The safety analysis was judged valid by all panel members and was identified by the clinical specialists as a salient finding requiring explicit consideration when interpreting the survival benefit. The colorectal surgeon noted that the increased serious-adverse-event signal was not prominent in their prior reading of the literature or in current guideline framing, and this interpretation was supported after joint review with the evidence-synthesis methodologists. The two colorectal oncologists likewise emphasized that toxicity represented a central trade-off in interpreting the survival benefit.

Panel feedback led to qualification of the conclusion wording to emphasize that the findings support preoperative systemic chemotherapy in appropriately selected patients while highlighting the accompanying toxicity cost that should guide patient selection.

## Data availability

OpenEBM-Corpus was reconstructed from publicly available studies. No private, proprietary, or patient-level data were used to construct the corpus. Source studies are identifiable by their public bibliographic records and registry identifiers where available. The reconstructed OpenEBM-Corpus, OpenEBM-Bench, evaluation datasets and associated metadata will be made available upon publication at https://github.com/zocskl/OpenEBM-Corpus. Data derived from third-party published sources are provided in accordance with the terms of the original sources.

## Code availability

All software used in this study is publicly available. All code related to this project can be accessed at https://github.com/zocskl/OpenEBM-Main.

## Acknowledgements

This study was funded by the GBRCE for Major Blinding Eye Diseases Prevention and Treatment, the Hainan Province Clinical Medical Center, the National Natural Science Foundation of China (82371086), and the Science and Technology Projects in Guangzhou (SL2024A03J00472; 2025A04J7150). Recruitment of domain experts in this study was supported by W.W.’s personal funds and resources.

## Author contributions statement

Study concept and design: W.W.; Acquisition, analyses, or interpretation: S.Y., J.W., H.X., Z.X.; Drafting of the manuscript: S.Y., W.W.; Critical revision of the manuscript for important intellectual content: S.Y., J.W., H.X., Z.X., W.W.; Statistical analyses: S.Y., J.W., H.X.; Obtained funding: W.W.; Administrative, technical, or material support: S.Y., J.W., W.W.; Study supervision: W.W.

